# CREST-GV: Cell types Ranking and Enrichment Score for selecTive Genetic Variants

**DOI:** 10.1101/2025.07.29.25332311

**Authors:** Simone G. Riva, Samvida S. Venkatesh, E. Ravza Gür, Nicolò Stranieri, Martin Sergeant, Matthew Baxter, Jim R. Hughes

## Abstract

Genome-Wide Association Studies (GWAS) have transformed our understanding of genetic traits and diseases since 2005 by analysing genetic variants across the human genome. Whilst they have successfully identified thousands of genetic locations linked to various conditions, converting these discoveries into practical understanding remains challenging. A key challenge lies in understanding through which cell types and tissues a specific set of genetic variants functions. This knowledge would aid researchers in designing experiments and identifying therapeutic targets. Causal variants often affect cell type specific *cis*-regulatory elements (CREs) and therefore the genetics of a trait are enriched within the CREs of effector cell types. Recent advances in genomics and computational methods are improving our understanding of genetic variants’ effects across cellular environments. In this work, we introduce Cell types Ranking and Enrichment Score for selecTive Genetic Variants (CREST-GV), a statistical tool that analyses genetic variants intersecting with open chromatin regions across hundreds of cell types, by performing a systematic computation of probability distributions and standardised metrics, transforming genomic data into quantitative enrichment measures with corresponding significance values. The tool demonstrates strong capability in prioritising disease-relevant cell types, for example, in diabetes studies where it correctly identified immune cells for Type 1 diabetes, and pancreatic islet cells and adipose tissue for Type 2 diabetes. CREST-GV shows robust generalisation across diverse genetic traits, successfully analysing variants associated with different conditions whilst maintaining consistent performance and adaptability through its flexible framework. This work includes a user-friendly web application connected to our public database of precomputed enrichment scores. The platform enables rapid querying across cell types and genetic variants, provides standardised results for comparative analysis, and allows the exploration of multiple cell type relationships simultaneously. This accessible interface makes complex enrichment analyses available to the broader research community, accelerating genetic research.

## 1 Introduction

Genome-Wide Association Studies (GWAS) have revolutionised our understanding of the genetic basis of complex traits and diseases by systematically examining genetic variants across the entire human genome. Since the first successful GWAS in 2005, this approach has identified thousands of genetic *loci* associated with various phenotypes, ranging from common diseases to quantitative traits [1]. However, despite the remarkable success in identifying disease-associated variants, translating these findings into functional insights remains a significant challenge [2, 3].

One of the most critical challenges in GWAS is understanding the cellular and tissue-specific context of identified genetic variants [4, 5]. Whilst GWAS can pinpoint genetic regions associated with a trait, they often struggle to elucidate the precise mechanisms by which these variants influence disease risk. Enrichment analyses have emerged as a powerful approach to address this limitation, aiming to determine which cell types and tissues the identified variants are most likely to exert their functional effects through [6, 7].

The complexity of enrichment studies lies in the heterogeneity of human tissues and cell types. Different cell types can have dramatically different gene expression profiles, epigenetic landscapes, and regulatory mechanisms. Traditional GWAS enrichment approaches face several challenges, for example: 1) limited reference data for comprehensive cell type coverage; 2) variability in tissue-specific gene regulation; 3) complex interactions between genetic variants and cellular contexts; and 4) technical limitations in capturing the full spectrum of cellular diversity.

Recent advances in single-cell genomics, functional genomics, and computational methods are helping to overcome these challenges, providing more nuanced insights into the cellular specificity of genetic associations. However, significant work remains to fully understand how genetic variants influence biological processes across different cellular environments [8, 9].

The landscape of GWAS enrichment analysis has evolved significantly in recent years, with sophisticated computational and statistical approaches emerging to decode the functional context of genetic variants. A landmark study by Finucane *et al*. (2015) introduced stratified linkage disequilibrium score regression, which revolutionised cell type specific enrichment analysis by quantifying heritability across different genomic annotations [10]. Building upon this foundation, specialised methods like CHEERS (Chromatin Element Enrichment Ranking by Specificity) have made substantial contributions to understanding variant functional relevance [11].

The CHEERS method, developed in the context of psychiatric disorders, represents a pivotal advancement in enrichment analysis. Specifically, the study “Genetic variants associated with psychiatric disorders are enriched at epigenetically active sites in lymphoid cells” demonstrated a novel approach to identifying cell type specific enrichment. By integrating epigenetic data from lymphoid cells with GWAS summary statistics, CHEERS provided insights into the potential functional mechanisms underlying psychiatric genetic risk.

In this work, we propose CREST-GV (Cell types Ranking and Enrichment Score for selecTive Genetic Variants), a statistical approach which allows querying our data collection of 457 cell types (as part of an ever expanding data collection) to determine the enrichment score of a set of genetic variants. CREST-GV relies on peaks, called open chromatin data, as a general proxy for regulatory activity, for each cell type in the data collection, assessed by leveraging LanceOtron peak caller [12]. To make it more appealing, CREST-GV can also query personal, or so-called *in-house* data.

## 2 Data & Method

The implementation of CREST-GV relies on an extensive collection of chromatin accessibility data and genetic variants. Our methodological approach encompasses three main components: (1) compilation and processing of open chromatin profiles across hundreds of cell types, (2) development of a statistical framework for enrichment analysis, and (3) implementation of computational procedures for score calculation and validation. This section details our data sources and analytical methods.

### 2.1 Data

We collected 457 different cell/tissue types from various samples and collections in genome assembly GRCh38 (hg38), allowing us to cover almost the entire human body. In detail, we selected 10 cell types from Turner *et al*. [13], 222 cell types from Zhang *et al*. [14] (equally divided between adult and fetal cells), 23 cell types from Granja *et al*. [15], 19 primary peripheral blood mononuclear cells (PBMC) cell types (See Table 1), 175 immune cell types from Calderon *et al*. [16], and 8 erythropoiesis cell types from Ludwig *et al*. [17]. Turner *et al*., Granja *et al*., and PBMCs datasets have been (upstream and downstream) analysed in more detail to isolate and generate bigWig files of the correct cell types of interest; Zhang *et al*. data has been taken from their interactive web atlas in bigWig format directly; whereas, Calderon *et al*. and Ludwig *et al*. datasets have been re-analysed by using CATCH-UP pipeline [18]. All 457 cell type bigWig tracks were peak called using LanceOtron [12], in order to identify significant open chromatin regions for downstream enrichment analysis. The collections are summarised in Table 2.

**Table 1.**
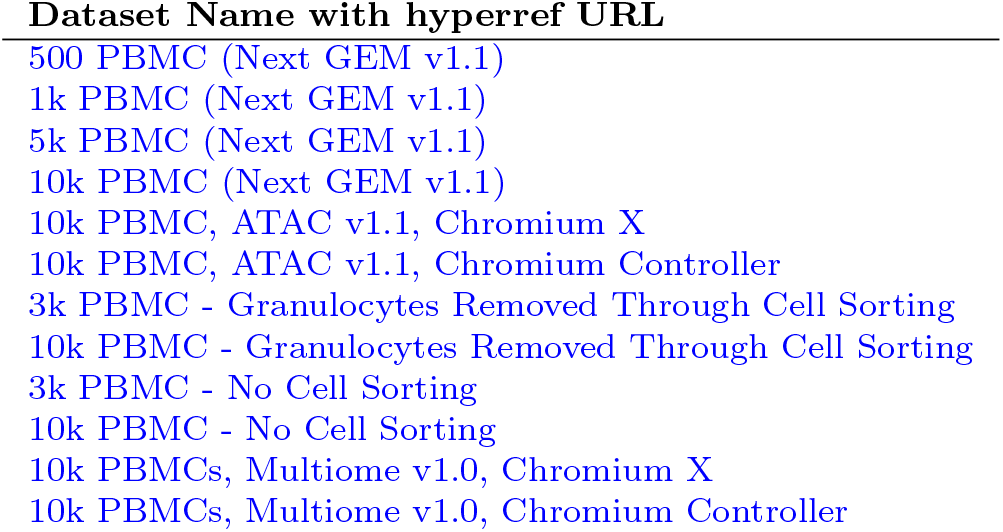
Human Healthy Donors, 10x Genomics, accessed 04/03/2023.

**Table 2.** Summary of data collections used in CREST-GV analysis. The table shows the collection names, number of distinct cell types in each collection, and corresponding source publications. Each collection represents a unique set of chromatin accessibility profiles from different cellular contexts.

| Collection name | Number different cell types | Authors |
| --- | --- | --- |
| cad | 10 | Turner *et al.* |
| catlas_fetal | 111 | Zhang *et al.* |
| catlas_adult | 111 | Zhang *et al.* |
| mpal | 23 | Granja *et al.* |
| super_pbmc | 19 | 10x Genomics |
| calderon | 175 | Calderon *et al.* |
| ludwig | 8 | Ludwig *et al.* |

### 2.2 Method

Consider the variants from the 1000 Genome Project data (*𝒢*) of size *G* [19], and *C*_*ij*_, a set of peak regions (*R*) specific for cluster/cell type *i* ∈ [1, …, *n*_*j*_], where *n*_*j*_ is the total number of clusters/cell types per collection *j* (collections are explained in Table 2). To note, disjoint peaks are found using LanceOtron, as stated in Section 2.1, on each coverage cluster/cell type specific sample. *𝒢* is intersected with 𝒞_*j*_ = {*C*_*ij*_, *i* ∈ [1, …, *n*_*j*_]}.

The intersection (number of a selected set *𝒳* of genetic variants falling inside *R*) and the corresponding proportion wrt the number of variants | *𝒳* |, respectively, are defined in eq. 1 and eq. 2.

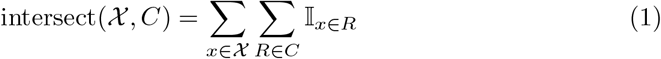

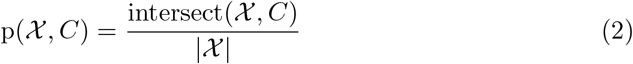

The Standard Error for specific cluster/cell type (SE_*ij*_), defined in eq. 4, quantifies the standard deviation of intersect (*𝒩, C*_*ij*_) given *𝒩* drawn with replacement from *𝒢*.

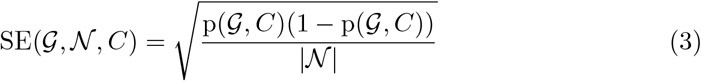

To measure the statistical significance and direction of the deviation from the expected proportion, the cluster/cell type specific Z-score is calculated using eq. 4, defining the enrichment score of the genetic *𝒩*.

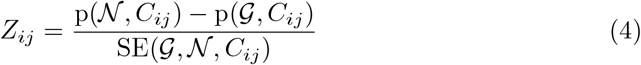

The *Z*_*ij*_ enrichment score is valid within each independent collection. To compare results across different collections, a standardisation approach has to be applied (eq. 5), creating a standard normal distribution.

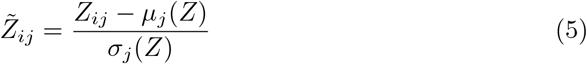

Now we have 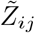 for each cluster/cell type *i*, within each collection *j*. Calculating its right-tailed *p*-value (eq. 6) allows us to quantify the probability of observing a proportion as large as or larger than the one obtained purely by chance.

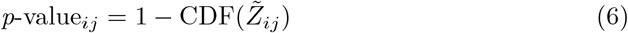

CDF or Cumulative Distribution Function is defined in eq. 7, which represents the probability of a random variable taking a value less than or equal to a specific point.

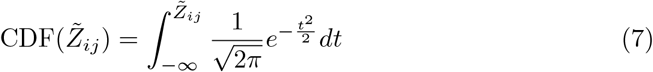

Saying so, using a significance threshold equal to 0.05, we can rank the enrichment score 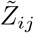, and also provide insight if 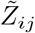 is statistically significant or not. Figure 1 provides a visual representation of the methodological workflow, illustrating the detailed steps described above.

**Figure 1.**
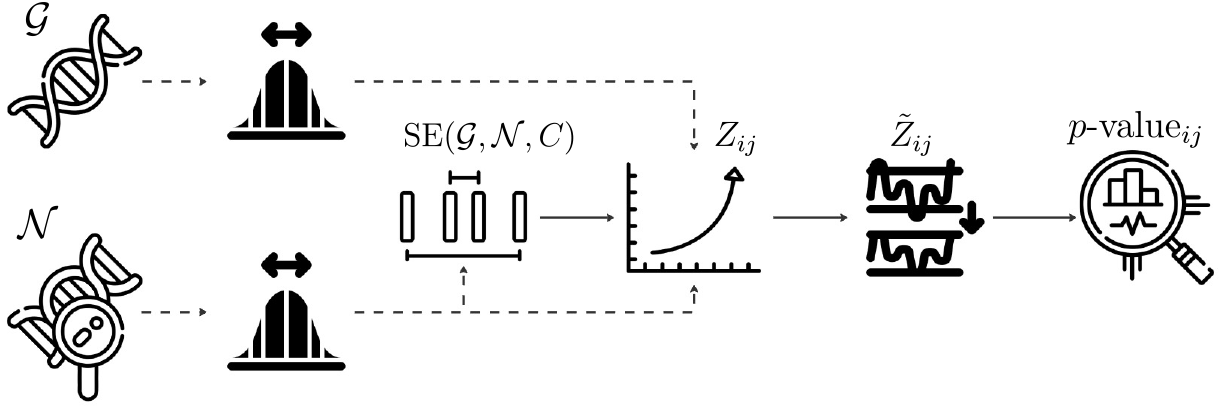
Schematic representation of the CREST-GV computational workflow. The pipeline begins with genetic variant input (*𝒢* and *𝒩*) and chromatin accessibility data, processes through statistical calculations including probability estimation, SE computation, Z-score calculation, normalisation, and concludes with significance assessment. Each icon represents a key computational step in the enrichment analysis process.

The statistical framework relies on the assumption of normality, which is verified when both |*𝒩*| *·*p(*𝒩, C*_*ij*_) and |*𝒩*| *·* (1 − p(*𝒩, C*_*ij*_)) exceed 5 for each cluster/cell type *i* and collection *j*. These conditions ensure the validity of our Z-score based statistical inference. When these conditions are met, we can reliably use the normal distribution for our statistical calculations, including Z-score computations and *p*-value assessments.

## 3 Results

The 1000 Genomes Project data (𝒢) serve as an ideal reference set for establishing baseline proportions of genetic variants overlapping with cell type specific regulatory elements. This choice is motivated by several key factors: (1) it represents a comprehensive and well-characterized collection of human genetic variation, (2) it is agnostic of any specific trait or disease, thus providing an unbiased background distribution, and (3) its large sample size and diverse population coverage ensure robust estimation of expected variant frequencies. Using this dataset as our null distribution enables us to accurately identify significant enrichments above the background level of genetic variation, making it particularly suitable for detecting cell type specific effects in trait-associated variants.

To demonstrate CREST-GV’s ability to identify disease/trait-relevant cell types through genetic variant enrichment, we analysed trait-associated variants from the GWAS Catalog [20], version 1.0.2, downloaded on 22/08/2023. To ensure comprehensive coverage of potentially causal regions, we expanded our analysis to include proxy SNPs in linkage (*R*^2^ ≥0.8) identified using TopLD [21] in the European population. This approach allows CREST-GV to evaluate the enrichment of both lead SNPs and their correlated variants across specific cell types.

Our initial dataset comprised 7, 778 trait-related GWAS studies from the GWAS Catalog. To maintain statistical robustness in our enrichment analyses, we established a minimum threshold of 500 variants per trait, post-imputation. This quality control step yielded a final set of 1, 125 trait-related GWAS, ensuring sufficient statistical power for detecting cell type specific enrichment signals.

To demonstrate the necessity of normalisation in our analysis, we compared the distribution of Z-scores across different data collections before and after standardisation. Raw scores (*Z*_*ij*_) often exhibited collection-specific biases and varying scales, making direct comparisons between different collections challenging. For instance, *Z*_*ij*_ from catlas adult collection ranges from −2 to 5, whilst calderon collection spans from 0 to 25 (see Figure 2), despite representing similar biological phenomena. After applying the normalisation procedure (equation 5), all collections showed comparable distributions with standardised ranges, enabling meaningful cross-collection comparisons, as shown in Figure 3.

**Figure 2.**
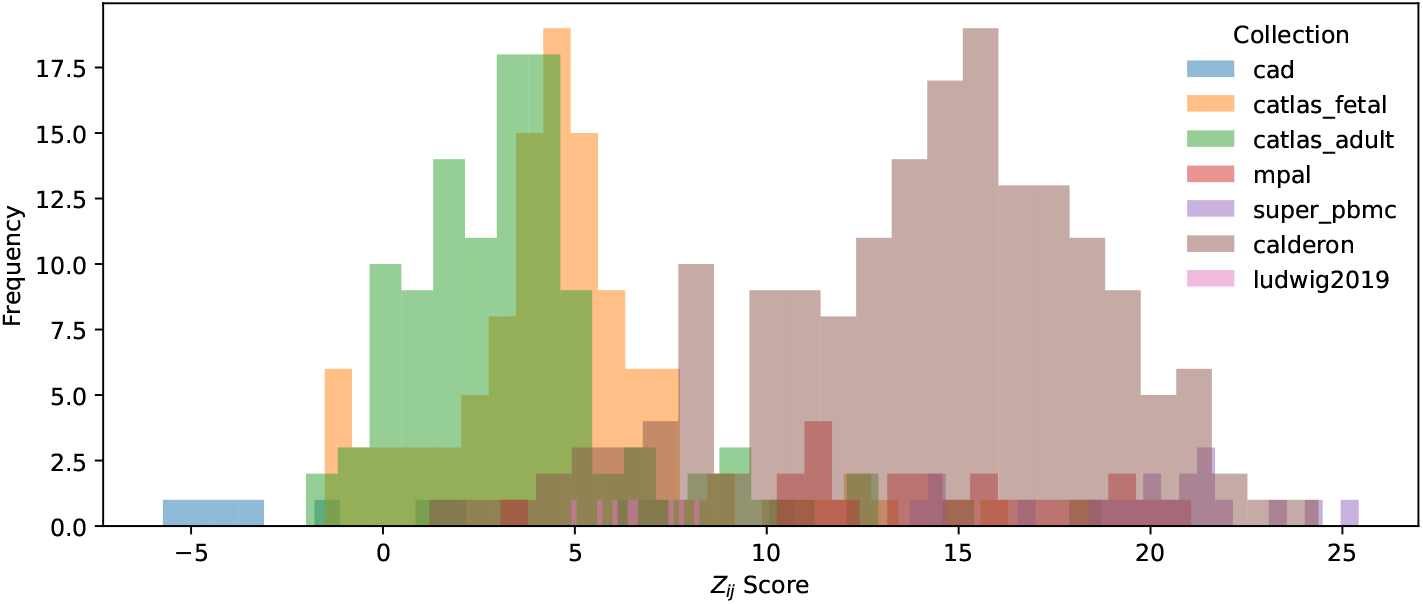
Raw *Z*_*ij*_ score distributions across different chromatin accessibility data collections. The histogram illustrates collection-specific variations in *Z*_*ij*_ score ranges and distributions, emphasising the necessity for normalisation in cross-collection analyses.

**Figure 3.**
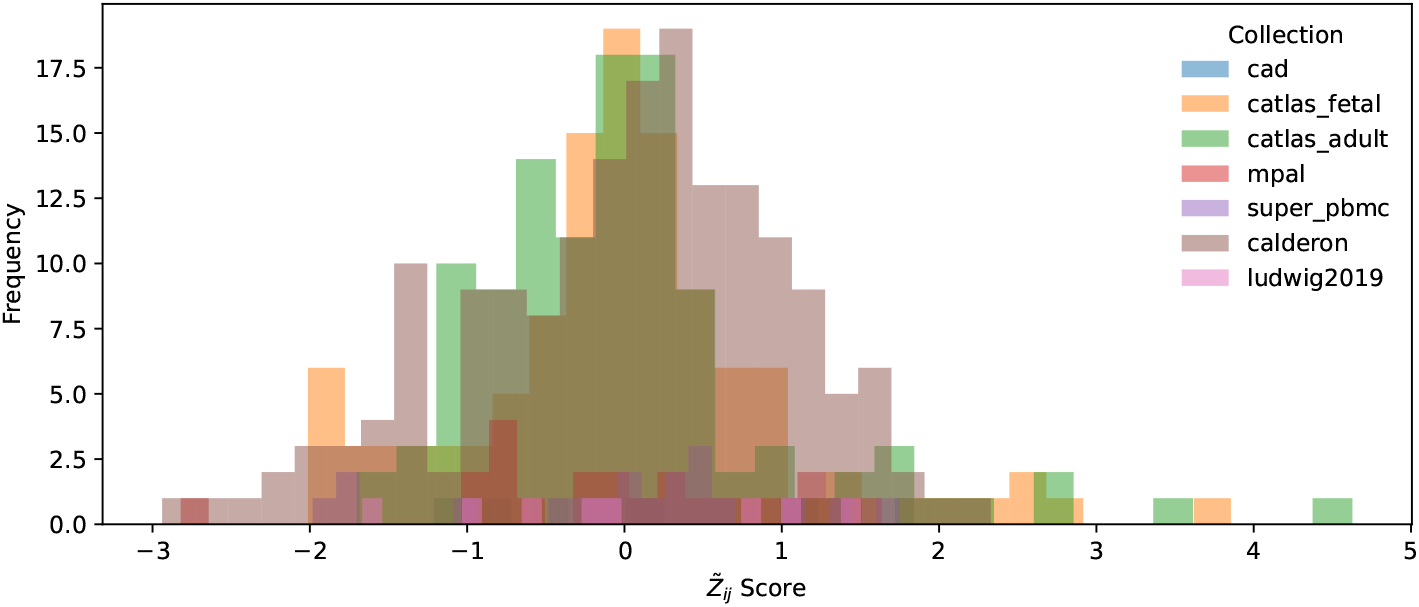
Standardised 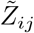 score distributions across chromatin accessibility data collections. After normalisation, all collections show comparable distributions centred around zero, facilitating meaningful cross-collection comparisons.

This standardisation proved particularly crucial when integrating results from multiple sources, ensuring that enrichment scores were evaluated on the same scale regardless of their origin, thus preventing any collection-specific biases from dominating the final cell type prioritisation.

To validate CREST-GV’s effectiveness, we investigated in detail 3 traits (out of the 1, 125 trait-related GWAS selected), dividing the analysis into 2 steps: (1) firstly, we analysed in detail Type 1 and Type 2 diabetes GWAS traits, two well-characterised diseases with distinct cellular mechanisms; (2) secondly, we looked closely at red blood cell trait distribution width, a haematological parameter known to be predominantly regulated in erythroid lineage cells. This combination of metabolic diseases and a cell type specific trait provides a robust test set for evaluating CREST-GV’s capacity to identify biologically relevant cell types across different physiological contexts.

Type 1 and Type 2 diabetes serve as ideal test cases due to their known cell type specificities: Type 1 diabetes involves autoimmune destruction of pancreatic beta cells, whilst Type 2 diabetes primarily affects metabolic tissues.

We analysed Type 1 and Type 2 diabetes GWAS data by combining enrichment scores across all collections and ranking cell types based on their normalised CREST-GV scores 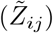. The analysis pipeline aggregated collection-specific results and identified the top and bottom 10 enrichment values, revealing distinct cell type signatures characteristic of each diabetes type’s underlying biology. The distinct cellular mechanisms underlying Type 1 and Type 2 diabetes are reflected in their CREST-GV enrichment patterns (Figure 4). For Type 1 diabetes (Figure 4a), the highest enrichment scores were observed in immune cell types, particularly T lymphocytes and CD4-positive cells, aligning with its known autoimmune pathogenesis. In contrast, Type 2 diabetes (Figure 4b) showed the strongest enrichment in pancreatic islet cells and adipose tissue, consistent with its metabolic nature. Notably, both diseases showed negative enrichment scores in cell types not directly involved in their pathogenesis, such as various neural cell populations, demonstrating CREST-GV’s ability to not only identify relevant cell types but also exclude biologically unrelated ones.

**Figure 4.**
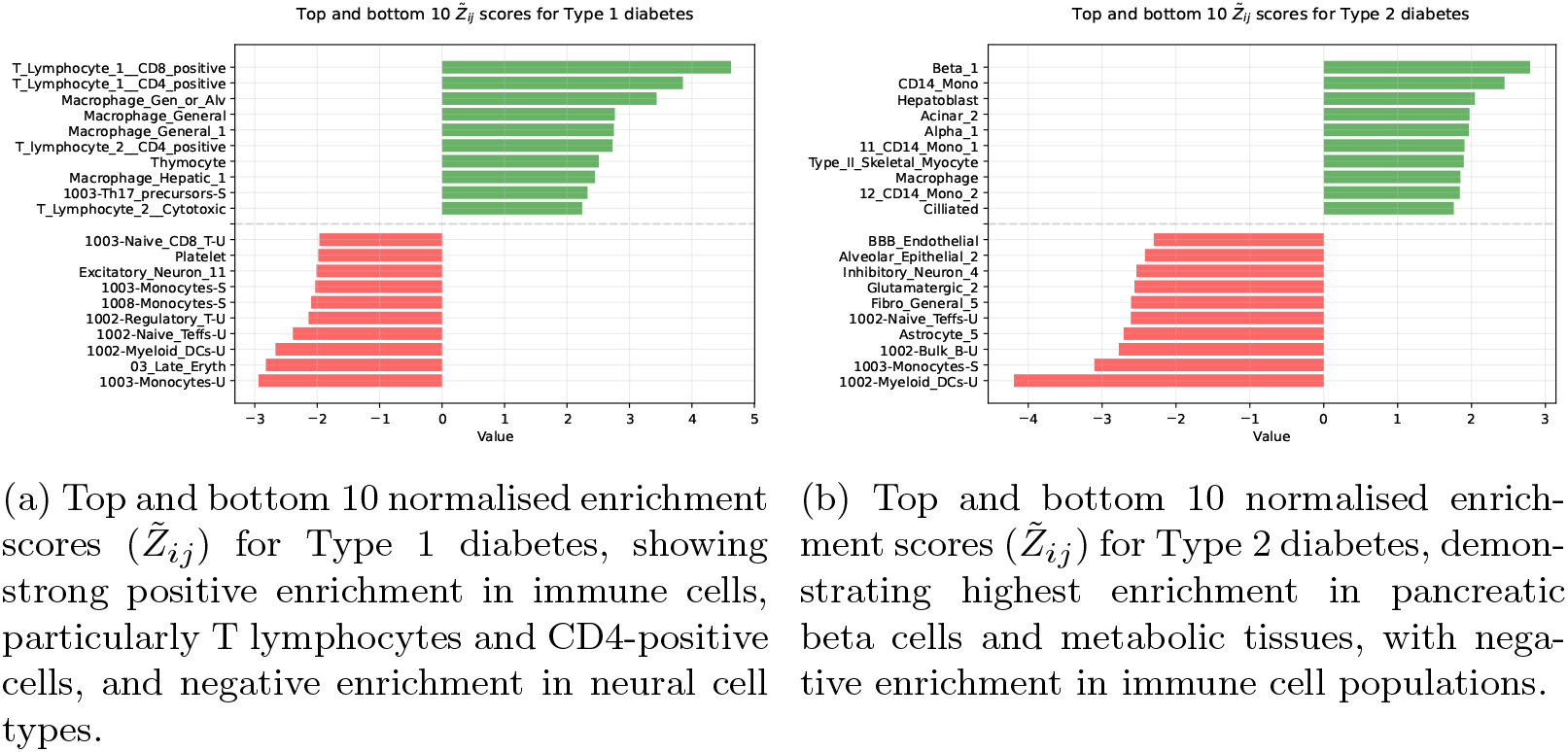
Comparison of cell type enrichment patterns between Type 1 and Type 2 diabetes. (a) Type 1 diabetes shows characteristic enrichment in immune cell populations, reflecting its autoimmune pathogenesis. (b) Type 2 diabetes is characterised by enrichment in metabolic tissues, particularly pancreatic cells, consistent with its metabolic nature. The green and red bars represent positive and negative enrichment scores, respectively.

To provide a comprehensive view of all significantly enriched cell types (*p*-value *<* 0.05) across collections, we generated word clouds where the size of each cell type label is proportional to its enrichment score (Figure 5). For Type 1 diabetes (Figure 5a), the visualisation prominently features T lymphocytes, CD4-positive cells, and various immune cell populations, with “CD14 Mono” and other immune-related cell types appearing prominently. In contrast, the Type 2 diabetes word cloud (Figure 5b) highlights “Beta_1”, “Alpha_1”, and “hepatoblast” among the most prominent cell types, reflecting the disease’s metabolic nature and the involvement of pancreatic islet cells and liver tissue. These visualisations effectively capture the distinct cellular landscapes of both diseases, reinforcing the findings from our ranked analysis.

**Figure 5.**
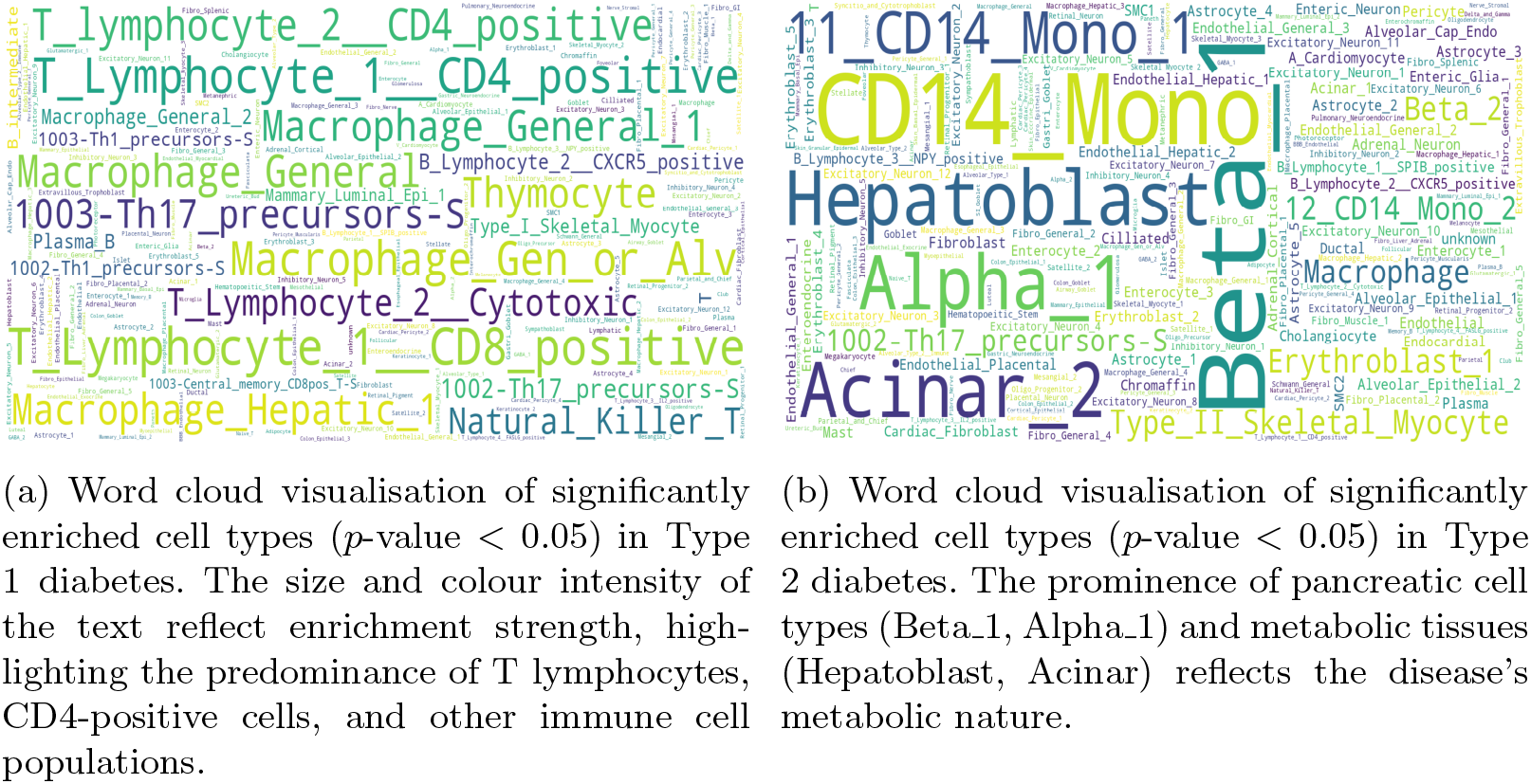
Word cloud representations of significantly enriched cell types in diabetes. (a) Type 1 diabetes enrichment pattern emphasises immune cell populations, particularly T lymphocytes and CD4-positive cells. (b) Type 2 diabetes shows predominant enrichment in pancreatic and metabolic cell types. Text size and colour intensity correspond to enrichment strength (*p*-value *<* 0.05).

As an additional validation of CREST-GV’s capability to identify biologically relevant cell types, we analysed genetic variants associated with red blood cell distribution width. Figure 6 presents compelling evidence of CREST-GV’s precision in cell type prioritisation. The barplot (Figure 6a) shows the highest enrichment scores in erythroid lineage cells, with “Erythroblast” populations showing the strongest positive enrichment, whilst neural and immune cell types display negative enrichment. This cell type specificity is further reinforced in the word cloud visualisation (Figure 6b), where “Erythroblast” populations and “03_Late_Eryth” appear prominently, alongside other hematopoietic cell types. The presence of both fetal and adult erythroid populations among the top enriched cell types reflects the developmental continuity of red blood cell production and the known large degree of regulatory overlap in the transcriptional control of these programmes.

**Figure 6.**
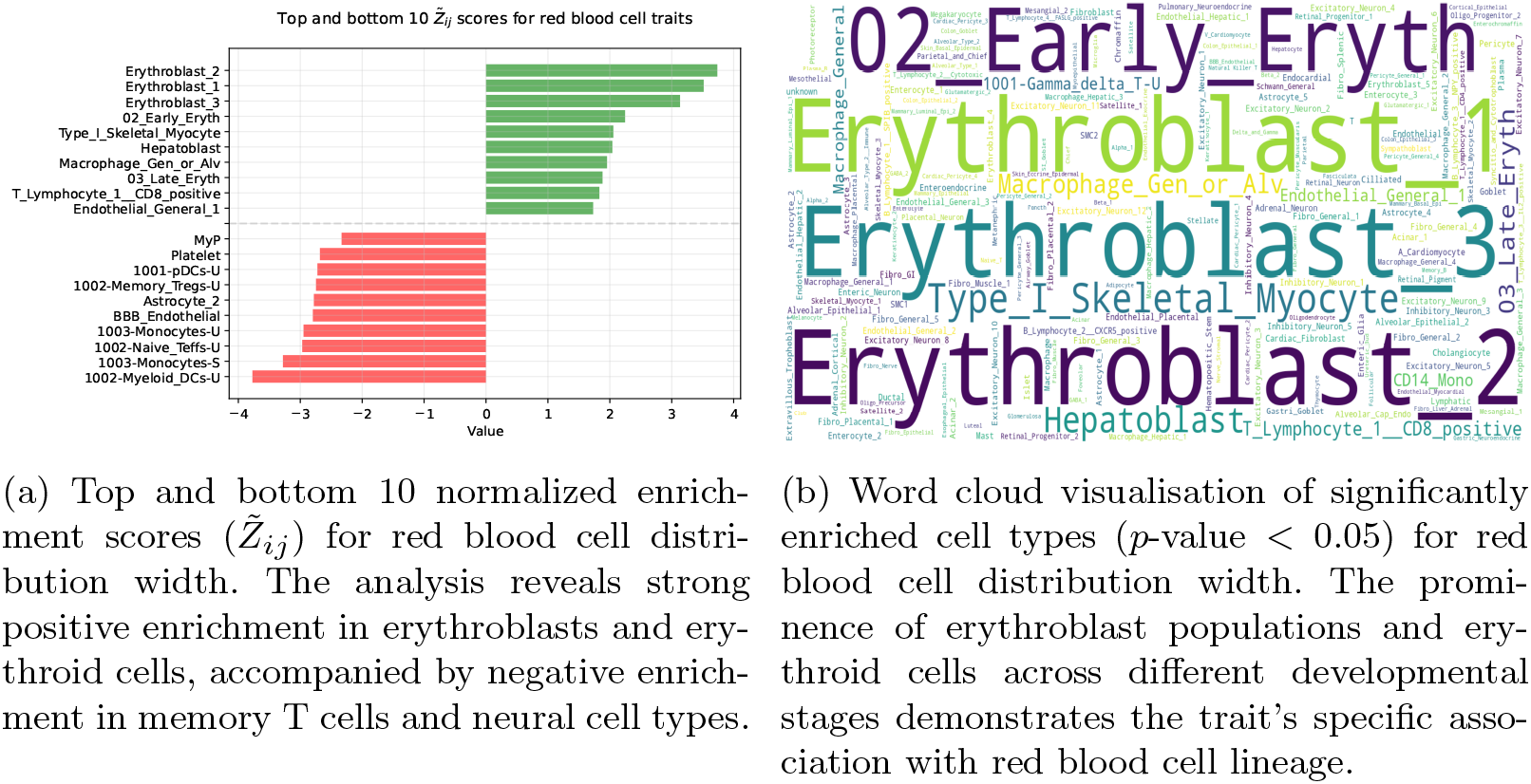
Cell type enrichment analysis for red blood cell distribution width. (a) Bar plot showing strong enrichment in erythroid lineage cells and negative enrichment in unrelated cell types. (b) Word cloud representation highlights the predominance of erythroblasts and erythroid cells, with varying contributions from different developmental stages and collections. Text size and colour intensity correspond to enrichment strength (*p*-value *<* 0.05).

## 4 Conclusion & future work

Understanding the target cell type of genetic variants is crucial for deciphering disease mechanisms and developing targeted therapies. Here we summarise CREST-GV’s contributions to this challenge and outline future developments to enhance its utility for the research community.

### 4.1 Conclusion

In this work, we presented CREST-GV (Cell types Ranking and Enrichment Score for selecTive Genetic Variants), a statistical framework developed to identify effector cell type in genetic variants. The tool analyses the intersection between trait-associated variants and chromatin accessibility profiles across hundreds of cell types, implementing a robust normalisation procedure to enable cross-collection comparisons. Using the 1000 Genome Project data as an unbiased reference, CREST-GV calculates enrichment scores and their statistical significance, effectively prioritising disease-relevant cell types. The method’s effectiveness was demonstrated through the analysis of three distinct traits: Type 1 diabetes, where it identified immune cell enrichment; Type 2 diabetes, highlighting pancreatic and metabolic tissues; and red blood cell traits, showing specific enrichment in erythroid lineages.

In detail, our analysis demonstrates CREST-GV’s effectiveness across diverse trait types. For Type 1 diabetes, we observed strong enrichment in immune cell populations, particularly T lymphocytes and CD4-positive cells, whilst non-immune cells showed negative enrichment, aligning with its autoimmune pathogenesis (Figure 4a). Type 2 diabetes revealed a distinct pattern with the highest enrichment in metabolic tissues, notably pancreatic beta and alpha cells, along with hepatoblasts, reflecting its metabolic nature (Figure 4b). The word clouds further reinforced these patterns, showing broader immune cell involvement in Type 1 diabetes (Figure 5a) and metabolic tissue enrichment in Type 2 diabetes (Figure 5b). As a validation case, analysis of red blood cell distribution width showed precise cell type specificity, with strongest enrichment in erythroblasts and late erythroid cells, whilst neural and immune cells displayed negative enrichment (Figure 6a). The corresponding word cloud highlighted both fetal and adult erythroid populations, along with related hematopoietic cell types (Figure 6b). These results collectively demonstrate CREST-GV’s ability to accurately identify disease-relevant cell types across different biological contexts, from autoimmune and metabolic diseases to traits with highly specific cellular origins.

### 4.2 Future work

The next phase of CREST-GV’s development will focus on two main directions: data expansion and interface implementation. We plan to continuously integrate new cell type specific chromatin accessibility profiles from emerging collections, expanding our current database of over 450 cell types. This ongoing data collection effort will provide increasingly comprehensive coverage of human cell types and developmental stages. A key objective is the development of a user-friendly web interface that will offer advanced querying capabilities, allowing users to explore precomputed enrichment scores across multiple traits simultaneously, perform comparative analyses between related phenotypes, and visualise results through interactive plots. Additionally, we aim to implement features for custom analysis submission, enabling researchers to analyse their own GWAS data against our growing cell type collection. These developments will strengthen CREST-GV’s utility as a resource for understanding the cellular basis of genetic traits and diseases.

## Data Availability

All data produced in the present work are contained in the manuscript.

## Acknowledgments

S.G.R. is supported by the MRC grant (MC UU 00029/3). S.S.V. is supported by Schmidt Sciences, LLC. E.R.G. is supported by the Ministry of National Education Selection and Placement of Candidates Sent Abroad for Postgraduate Education (YLSY) scholarship, Republic of Türkiye Ministry of National Education and the Wellcome Trust grant (225220/Z/22/Z). M.S, and M.B. are supported by the Wellcome Trust grant (225220/Z/22/Z). J.R.H. is supported by the Wellcome Trust grants (225220/Z/22/Z and 106130/Z/14/Z) and the MRC grant (MC UU 00029/3).

## Declaration

J.R.H. is a co-founder and director of Nucleome Therapeutics and provides consultancy to the company.

